# “Blame the bats again”: YouTube reactions to the 2026 Ebola public health emergency reveal mistrust, misinformation, and geopolitical anxiety

**DOI:** 10.64898/2026.07.22.26358723

**Authors:** Kaweesi Calvin Nantalaga, Alice Nantege

## Abstract

The World Health Organization declared the 2026 outbreak of Ebola disease caused by Bundibugyo virus in the Democratic Republic of the Congo and Uganda a Public Health Emergency of International Concern. Public engagement on video-sharing platforms such as YouTube offers insight into public perceptions during such crises, particularly in the affected region, yet these reactions remain largely uncharacterised. We analysed public discourse and sentiment surrounding the outbreak, focusing on thematic trends in YouTube comments, using a qualitative synthesis that combined thematic content analysis with topic modelling. Videos were identified through the YouTube Data application programming interface, which returned 50 videos each for the search terms “Ebola” and “Ebola virus disease.” After removing duplicates and excluding videos published before the 17 May 2026 emergency declaration, the 10 most-viewed videos were retained. From 4,457 extracted comments, 4,087 were analysed using Latent Dirichlet Allocation topic modelling, complemented by a lexicon-based sentiment analysis. Five themes emerged: geopolitical and cultural context; public figures, conspiracy theories, and misinformation; disease spread and transmission; public health measures and preparedness; and religious and spiritual interpretation. Discourse was strongly shaped by border concerns, mistrust of institutions, global aid politics, and comparisons with COVID-19, and sentiment was split near-evenly between fear and trust. Effective health communication during this outbreak must therefore address not only the scientific and medical dimensions of the emergency but also its geopolitical, cultural, and religious dimensions, while countering misinformation and building public trust.

## Introduction

Ebola virus disease is a severe, often fatal zoonotic illness that has repeatedly tested public health systems across sub-Saharan Africa. Caused by orthoebolaviruses and transmitted through direct contact with the body fluids of infected people or animals, it is characterised by high case fatality and a capacity to spread rapidly where surveillance, health infrastructure, and community trust are strained. On 17 May 2026, the World Health Organization (WHO) declared the outbreak of Ebola disease caused by Bundibugyo virus in the Democratic Republic of the Congo (DRC) and Uganda a Public Health Emergency of International Concern (PHEIC) [1]. The declaration followed a rapidly expanding cluster of cases in Ituri Province, with cross-border spread into Uganda, and was issued for a virus species against which no vaccine or treatment of proven efficacy is currently available [2].

Outbreaks of this kind unfold not only in clinics and communities but also across digital platforms, where the public interprets, debates, and reacts to unfolding events in real time. During health emergencies, social media both reflects and amplifies public sentiment, shaping how official guidance is received and whether it is trusted [3]. The World Health Organization has characterised this dynamic as an infodemic, an overabundance of information, accurate or otherwise, that can distort risk perception, erode institutional trust, and impede the uptake of protective measures [4]. Analyses of recent emergencies, including the 2022 mpox outbreak, have shown that video-sharing and social media platforms carry large volumes of both credible information and misinformation, and that outbreak discourse is frequently politicised and stigmatising [5]. Understanding this discourse is therefore central to effective risk communication, particularly in a region where the outbreak is not a distant event but an immediate concern for local populations. Yet public reactions to the current Ebola emergency remain largely uncharacterised, and YouTube, one of the most widely used video-sharing platforms, has received little attention in this context.

In this study, we analysed public discourse and sentiment surrounding the 2026 Ebola outbreak, focusing on thematic trends in YouTube comments. By examining how viewers engaged with widely viewed content in the weeks following the PHEIC declaration, we sought to identify the dominant themes shaping public perception and to draw out their implications for health communication during the ongoing response.

## Materials and methods

### Study design and video selection

We identified relevant videos using the search terms “Ebola” and “Ebola virus disease” through the YouTube Data API in R. For each term, the API returned 50 videos sorted by YouTube’s default “relevance” ranking. From the 100 videos returned, 12 duplicates appearing under both terms were removed, and videos uploaded before 17 May 2026, the date on which the WHO declared the Bundibugyo virus disease outbreak a Public Health Emergency of International Concern, were excluded. The remaining videos were ranked by view count, and the 10 most-viewed were selected for detailed analysis. This ensured that the analysed videos represented highly engaged content relevant to the outbreak during the study period.

### Comment extraction and topic modelling

We then retrieved all available top-level comments from these videos on 13 June 2026. Comments were disabled on one of the 10 videos; the remaining nine videos contributed the analysed comments. Comments were extracted using the YouTube Data API and processed in R. Text was normalised by converting to lower case, removing URLs and non-alphabetic characters, and collapsing whitespace; comments containing fewer than three words after cleaning were discarded, and English stop words were removed. A document-term matrix was constructed from the remaining tokens, excluding terms that occurred fewer than twice, leaving 4,087 comments for analysis. A 20-topic Latent Dirichlet Allocation (LDA) model [6] was then fitted using the text2vec package [7], which implements LDA through the WarpLDA sampler, with a document-topic prior of 0.1, a topic-word prior of 0.01, 2,000 iterations, and a fixed random seed (1234) for reproducibility. For each topic, we identified the top terms and used these to derive overarching themes and subthemes, which were refined according to relevance and frequency. Each comment was assigned to its dominant topic, and topics were grouped into five themes; the reported theme percentages reflect the proportion of comments whose dominant topic fell within each theme (S2 Table). Representative comments were selected for their alignment with the identified themes and cross-checked against thematic trends to ensure they reflected prevalent sentiments.

### Sentiment analysis

To complement the thematic analysis, we conducted a lexicon-based sentiment analysis of the retained comments using the NRC emotion lexicon [8] implemented in the syuzhet package, classifying comments by overall valence and by discrete emotional categories. This allowed quantification of the affective tone accompanying the dominant themes.

### Software

All analyses were conducted in R version 4.5.3. Video and comment retrieval used the httr and jsonlite packages; text preprocessing used the tm and tidytext packages; the 20-topic Latent Dirichlet Allocation model was fitted using text2vec version 0.6.6 (Warp-LDA, collapsed Gibbs sampling); and sentiment was assessed with the NRC Emotion Lexicon via the syuzhet package version 1.0.7.

### Reporting and ethics

We followed the STROBE guidelines in reporting this study [9]. No ethical review was required, as the study used publicly available data and involved no human subjects; comments were anonymised, no usernames or identifying details were reproduced, and quotations were lightly edited where necessary to prevent traceability.

## Results

From the qualitative synthesis of these videos, 4,087 meaningful comments were analysed, and five major themes emerged (Table 1).

**Table 1.** Distribution of the 4,087 analysed comments across the five themes.

| **Theme** | **Comments (n)** | **%** |
| --- | --- | --- |
| Public figures, conspiracy theories, and misinformation | 1,164 | 28.5 |
| Geopolitical and cultural context | 1,043 | 25.5 |
| Disease spread and transmission | 880 | 21.5 |
| Public health measures and preparedness | 825 | 20.2 |
| Religious and spiritual interpretation | 175 | 4.3 |
| **Total** | **4,087** | **100** |

### Geopolitical and cultural context

This theme accounted for 25.5% of comments. The Borders and Travel subtheme captured strong demands for border and flight restrictions with affected countries, while the Regional Focus subtheme reflected discussion of the DRC and the wider African region as the outbreak’s epicentre, frequently framed against global power and resource interests. One commenter questioned the geopolitical framing, asking “how the whole narrative would change if Congo had no exploitable minerals,” while another pushed back on generalisations about the continent, noting that “not all countries in Africa have Ebola. Africa is made of many countries.”

### Public figures, conspiracy theories, and misinformation

This was the most prominent theme, accounting for 28.5% of comments, centred on distrust of institutions and unfounded claims. The Conspiracy subtheme included assertions that “viruses are made in a laboratory” and that “African leaders have trusted the West too much,” while the Misinformation subtheme captured accusations of orchestrated alarm, with one user insisting they would not “fall for those lies anymore… fearmongering posters, videos, lies.” Discussion also linked the outbreak to international aid decisions and named public figures, reflecting broader scepticism toward global health institutions.

### Disease spread and transmission

This theme accounted for 21.5% of comments and addressed how Ebola spreads and how dangerous it might become. A Transmission Routes subtheme featured discussion of animal sources and bushmeat, including resigned remarks such as “it’s only confined until it’s not. Oh, blame the bats again.” A Comparative subtheme drew repeated parallels with COVID-19 and other outbreaks, with one commenter listing “measles, hanta, Ebola, COVID” and concluding that “humanity is not prepared.”

### Public health measures and preparedness

This theme accounted for 20.2% of comments and covered vaccines, protective behaviour, and health-system capacity. The Vaccination subtheme reflected frustration over the pace of development, exemplified by the question, “COVID came out and within a year we had vaccines, but Ebola has been a threat for years and still no vaccine?” The Preparedness subtheme captured practical concerns about containment, with one user observing that “finding people who have been exposed is extremely difficult since people are always travelling around the world.”

### Religious and spiritual interpretation

This theme accounted for 4.3% of comments. Commenters framed the outbreak in spiritual terms, debating divine versus human causation, “is it the government or is it God doing this?”, and expressing faith-based responses such as “I hope and pray that it will not spread worldwide.”

### Sentiment

Lexicon-based analysis showed a near-even split between negative (4,390) and positive (4,287) valence. Among discrete emotions, fear (2,846) and trust (2,841) were the most prominent and almost equally weighted, followed by anticipation (2,226) and sadness (2,201), and then anger (1,720), joy (1,483), disgust (1,483), and surprise (1,362) (Fig 1). This balance indicates that public discourse was shaped simultaneously by apprehension about the outbreak and by expressions of trust and hope.

**Fig 1.**
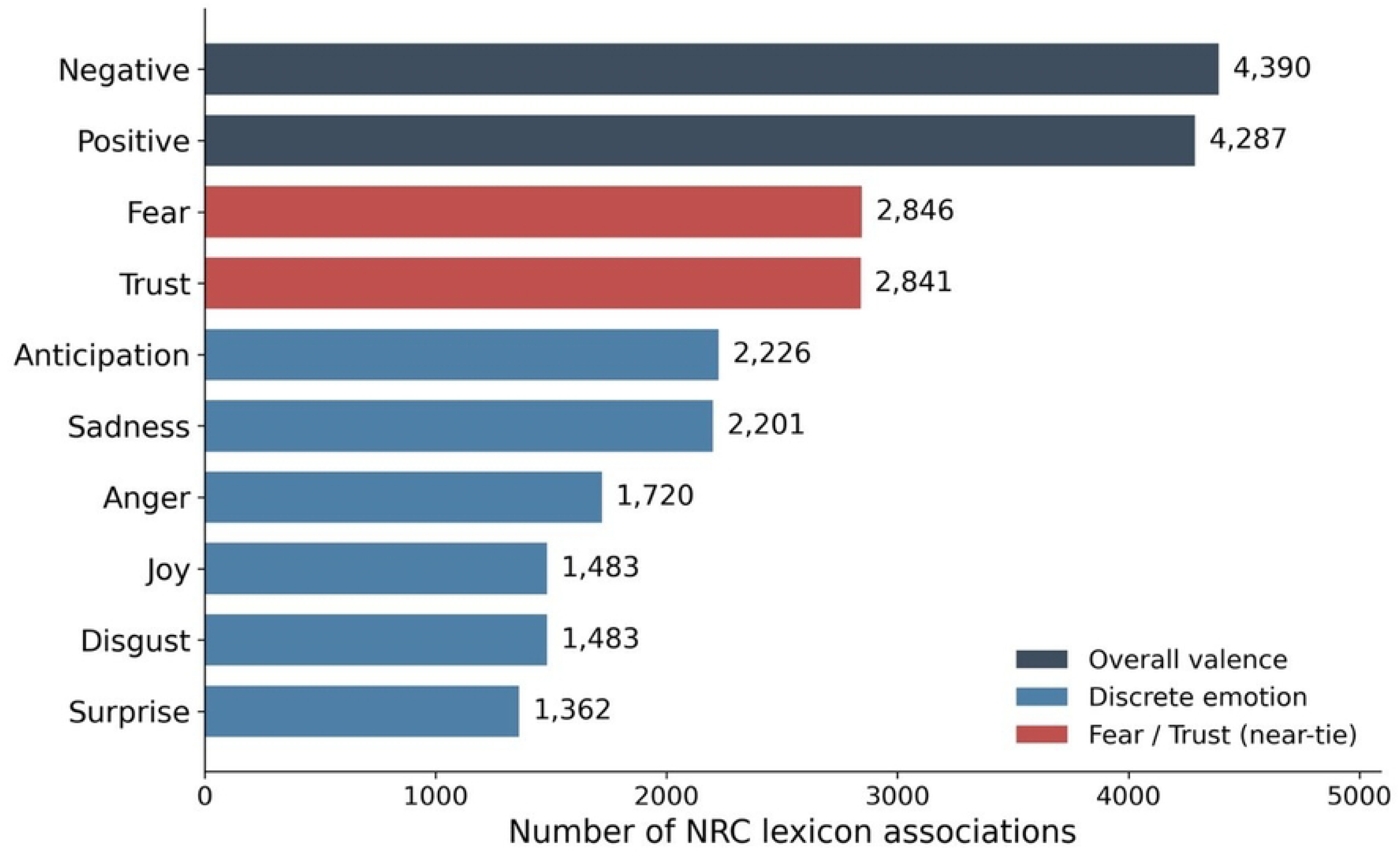
NRC sentiment and emotion profile of the analysed comments. Bars show the total number of NRC Emotion Lexicon associations for the two overall valence categories and the eight discrete emotions across the 4,087 analysed comments; a single comment may contribute associations to more than one category.

## Discussion

This analysis highlights the role digital platforms play in reflecting and amplifying public sentiment during a major health emergency unfolding within our own region. As a Public Health Emergency of International Concern affecting the DRC and neighbouring Uganda, the 2026 Bundibugyo Ebola outbreak is not a distant event but an immediate regional concern, and YouTube discourse offers a window into how publics interpret and respond to it in real time. The prominence of borders, travel, and regional framing emphasises the perceived transnational nature of the outbreak and public anxiety about its international spread. The repeated characterisation of the DRC and Africa as the epicentre reflected both genuine awareness and underlying bias, with some commenters reducing a diverse continent to a single source of contagion while others actively contested this framing. Public health messaging must therefore address not only the medical dimensions of the outbreak but also the geopolitical fears and prejudices that shape how information is received.

A distinctive feature of this discourse was the intertwining of the outbreak with global aid politics and named public figures, including discussion of reductions in international health funding and their perceived consequences for the response. Whereas analyses of earlier emergencies such as the 2022 mpox outbreak found politics to be a comparatively minor strand of online discourse [10], the Ebola discourse examined here was pervaded by references to aid decisions and named political figures, suggesting that contemporary public engagement with health emergencies is increasingly filtered through partisan and geopolitical lenses. Coupled with the predominance of conspiracy theories and accusations of “fearmongering,” this underscores the challenge facing health communicators: mistrust of institutions can attach itself to an outbreak response regardless of its scientific merits, eroding the credibility of legitimate public health guidance [11].

The comparative framing of Ebola against COVID-19 was pervasive and double-edged. While it helped audiences contextualise an unfamiliar threat, it also imported the polarisation and fatigue of the pandemic, including scepticism toward vaccines, lockdowns, and authorities [12].

Discussion of bushmeat and animal sources, distinctive to Ebola, further reflected attempts to locate blame and assign responsibility, sometimes along cultural lines.

The sentiment profile is particularly instructive. The near-equal weighting of fear and trust suggests that public discourse was not uniformly hostile or alarmist; rather, apprehension coexisted with genuine confidence and hope. This balance represents an opportunity.

Communication strategies that acknowledge fear while actively reinforcing trust, through transparency, consistency, and engagement with affected communities, may resonate with an audience that is anxious but not yet disengaged. The strong thread of religious and spiritual interpretation reinforces the need for culturally sensitive messaging that works alongside, rather than against, community belief systems.

However, this study has several limitations. First, YouTube comments may not represent the full spectrum of public opinion, as engagement varies across demographic groups, potentially introducing bias. The discourse analysed was also predominantly in English and skewed toward international audiences, which may underrepresent the perspectives of populations within the DRC and Uganda most directly affected. Second, selecting the top 10 most-viewed videos emphasises highly visible content and may overlook significant videos with lower view counts. Third, reliance on comment analysis excludes viewers who did not comment. Fourth, the lexicon-based sentiment analysis infers emotion from individual word associations and cannot fully capture negation, sarcasm, or context, so the affective profile should be read as indicative rather than definitive. These limitations underscore the importance of interpreting our findings within the context of YouTube’s user base and engagement patterns.

## Conclusion

Amidst the ongoing Bundibugyo Ebola outbreak in the DRC and Uganda, public health communication must be holistic, addressing not only the scientific and medical aspects of the emergency but also the geopolitical, cultural, and religious dimensions that shape public perception. The near-equal balance of fear and trust in public discourse represents a genuine opportunity: messaging that acknowledges anxiety while actively building trust, countering misinformation, and engaging affected communities may prove most effective in sustaining public confidence during the response.

## Acknowledgments

None.

## Data availability

The data underlying this study are YouTube comments that are publicly viewable on YouTube but cannot be redistributed by the authors under the YouTube API Services Terms of Service, which prohibit the distribution of retrieved content to third parties. To support reproducibility, the two search terms and the identifiers of the 10 analysed videos are provided in S1 Table, and the R scripts used for topic modelling and sentiment analysis are available from the authors on reasonable request.

## Author contributions

Kaweesi Calvin Nantalaga: Conceptualization, Data curation, Formal analysis, Investigation, Methodology, Software, Writing - original draft, Writing - review and editing. Alice Nantege: Data curation, Investigation, Writing - review and editing.

## Supporting information

**S1 Table**. Search terms and identifiers of the 10 analysed YouTube videos.

**S2 Table**. Mapping of the 20 Latent Dirichlet Allocation topics to the five themes.

